# A prospective, single-center study to evaluate the clinical performance of Meril ABFind in individuals vaccinated against COVID-19

**DOI:** 10.1101/2022.01.15.22269231

**Authors:** Jayanthi Shastri, Sachee Agrawal, Nirjhar Chatterjee, Harsha Gupta

## Abstract

**Background:** Accurate rapid antibody detection kits requiring minimum infrastructure are beneficial in detecting post-vaccination antibodies in large populations. ChAdOx1-nCOV (COVISHIELD™) and BBV-152 (Covaxin™) vaccines are primarily used in India.

**Methods:** In this single-centre prospective study, performance of Meril ABFind was investigated by comparing with Abbott SARS-CoV-2 IgG II Quant (Abbott Quant), GenScript cPass SARS-CoV-2 neutralization antibody detection kit (GenScript cPass), and COVID Kawach MERILISA (MERILISA) in 62 vaccinated health care workers (HCW) and 40 pre-pandemic samples.

**Results:** In the vaccinated subjects, Meril ABFind kit displayed high sensitivity of 93.3% (CI, 89.83%-96.77%), 94.92% (CI, 91.88%-97.96%), and 90.3% (CI, 86.20%-94.4%) in comparison to Abbott Quant, MERILISA, and GenScript cPass respectively. The results of the Meril ABFind in the COVISHIELD-vaccinated group were excellent with 100% sensitivity in comparison to the other three kits. In the Covaxin-vaccinated group, Meril ABFind displayed sensitivity ranging from 80% to 88.9%. In control samples, there were no false positives detected by Meril ABFind, while Abbott Quant, MERILISA, and GenScript cPass reported 2.5%, 10.0%, and 12.5% false positives, respectively. In the pre-pandemic controls, specificity of Meril ABFind was 100%, Abbott Quant 97.5%, MERILISA 90%, and GenScript cPass 87.5%.

**Conclusion:** The Meril ABFind kit demonstrated satisfactory performance when compared with the three commercially available kits and was the only kit without false positives in the pre-pandemic samples. This makes it a viable option for rapid diagnosis of post vaccination antibodies.

## 1 Introduction

SARS-CoV-2 which belongs to Betacoronavirus genus and Coronaviridae family is the causative agent for COVID-19. The spike protein (S-glycoprotein) present on the surface of the virus has a receptor binding domain (RBD) which binds to the angiotensin-converting enzyme 2 (ACE2) present on the cell surfaces and mediates the entry of this virus inside the host cell (1). Within the first three weeks of getting infected with the virus, the patients develop antibodies against the RBD of the viral spike protein (2). Early development of antibodies is important for survival and control of infection in the patients. These neutralizing antibodies provide protection from future infection by interfering with the interaction between the virus’s spike RBD and host’s ACE2.

India has primarily used ChAdOx1-nCOV (COVISHIELD™) and BBV-152 (Covaxin™) vaccines with health care workers (HCW) being the first group to get vaccinated. Serological tests can detect the humoral immune response following past infection and vaccination, and they can be useful for serological studies. Serology has also been used to make clinical decision on therapeutic interventions (3). While accurate laboratory based serological assays are available in India, a large rural population mandates reliable serological assays to detect humoral immunity following vaccination and post infection which can be performed without expensive laboratory equipment and cumbersome techniques. To address this unmet need, several companies are working on the development of reliable antibody testing kits with adequate sensitivity and specificity (4). Different diagnostic kits are available to identify individuals with immune response after the vaccine administration. Testing a huge number of vaccinated individuals with a diagnostic assay which requires complicated instruments and infrastructure would be cumbersome and would not be economical. In the given situation, a rapid test would be advantageous owing to speed of performance, quick results and minor requirements of equipments and infrastructure.

The Meril ABFind kit (Meril Diagnostics Pvt. Ltd., India) can detect circulating SARS-CoV-2 neutralizing antibodies that block the interaction between the RBD of the viral spike glycoprotein and the ACE2 cell surface receptor in serum, plasma and whole blood. This study was designed to investigate the clinical performance of this rapid antibody testing kit by evaluating its diagnostic sensitivity and specificity for the qualitative detection of neutralizing antibodies in defined and characterized specimens. For evaluating the performance of this kit, the results obtained were compared with the results obtained using three other commercially available kits viz. Abbott SARS-CoV-2 IgG II Quant (Abbott Diagnostic, USA), GenScript cPass SARS-CoV-2 neutralization antibody detection Kit (GenScript, USA), and COVID Kawach MERILISA (Meril Diagnostic, Pvt. Ltd., India).

## 2 Materials & Methods

### 2.1 Study Participants

A total of 62 participants of which 32 had been vaccinated with COVISHIELD and 30 with Covaxin, were recruited for this study in the months of June, July and August 2021. All recruited participants were health care workers working in the city of Mumbai, India. The inclusion criteria for the study were: healthcare workers in the age range of 18 to 60 years, fully vaccinated, i.e., two weeks after the second dose of COVISHIELD or Covaxin and less than 12 weeks after the second dose of COVISHIELD or Covaxin. Any individual who reported symptoms suggestive of COVID-19 were to be excluded, however none of the participants showed any such symptoms. The study was approved by the Institutional Review Board of Kasturba Hospital of Infectious Diseases, Mumbai, India; IRB number 05/2021. Written informed consent was obtained from all study participants. All collection, processing of samples and archiving of results were performed under approval from the Institutional Review Board.

The baseline demographics and the medical history of the participants were collected for analysis and evaluation.

### 2.2 Sample collection

One 5 ml vial of blood was collected by venipuncture from the cubital vein in serum separator tubes. Separated plasma was used for testing in all four tests. 40 stored pre-pandemic samples were used as a known negative control. These samples were collected in 2018 from patients with an acute febrile illness. Plasma was separated and stored at -80 °C in our laboratory (Molecular Laboratory, Kasturba Hospital for Infectious Diseases, Mumbai, India).

### 2.3 Serological Tests

All the samples were tested first using Abbott SARS-CoV-2 IgG II Quant (Abbott Quant). Then the samples were tested using the Meril ABFind kit and the two commercially available kits, COVID Kawach MERILISA (MERILISA) and GenScript cPass SARS-CoV-2 neutralization antibody detection kit (GenScript cPass).

The Abbott Quant kit results were beneficial in comparing anti-RBG IgG in the COVISHIELD- and Covaxin-vaccinated subjects in a quantitative manner. Meril ABFind kit is a qualitative membrane-based immunoassay for the detection of SARS-CoV-2 neutralizing antibody in whole blood, serum or plasma. This detection kit utilizes a colloidal gold labeled SARS-CoV-2 recombinant Receptor Binding Domain (rRBD) which binds with the SARS-CoV-2 neutralizing antibody of IgG class if present in the sample to give a colored band. A control band must be present for the test to be considered valid. A control band and a test band was considered positive, a control band without a test band was considered negative and absence of the control band was considered invalid irrespective of the test band. The test specimen was diluted in 1:10 ratio with sample dilution buffer and was added on the membrane strip, the results were read at the end of 20 minutes.

The Abbott SARS-CoV-2 IgG II Quant assay is a two-step automated chemi-luminescent microparticle immunoassay (CMIA) intended for the qualitative and quantitative determination of IgG antibodies to the RBD of the spike protein of SARS-CoV-2 in human serum and plasma. The analytical measurement interval was stated as 21 to 40,000 AU/ml, and positivity cut off was ≥50 AU/ml (defined by manufacturer) (5).

GenScript cPass SARS-CoV-2 neutralization antibody detection kit is a blocking ELISA detection tool which mimics the virus neutralization process. It uses horseradish peroxidase (HRP) conjugated recombinant RBD protein and human angiotensin converting enzyme-2 (ACE-2) receptor protein. The protein interaction between HRP-RBD and ACE-2 can be blocked by neutralizing antibodies against SARS-CoV-2 RBD. Signal inhibition was calculated as follow:

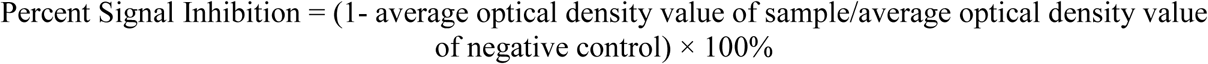

The test results were interpreted as positive when the percent signal inhibition was ≥30%, which was the cut-off for signal inhibition claimed by the manufacturer (6).

Covid Kawach MERILISA is an immune assay based on indirect enzyme linked immunosorbent assay principle for qualitative detection of IgG antibodies against SARS-CoV-2 in the sample. The optical density (OD) of the plate is read at 450 nm. This kit comes with a positive and negative control which works as marker of the kit. P/N ratio of positive control is defined as ratio of OD value of positive control to the average of the ODs of negative control. The test is considered to be valid if P/N ratio of positive control is greater than 1.5.

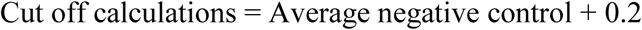

For the test sample, if OD value was greater than the cut-off value and P/N ratio was more than 1.5, sample was considered positive. While, for the test sample, if OD value was less than the cut-off value and P/N ratio was less than 1.5, sample was considered negative (7).

### 2.4 Managing Test Results

Results of all four tests were given to participants with the understanding that testing was done in the context of a research study. No clinical decisions were made on the basis of this study. Participants were informed that a positive result did not preclude COVID-19 in the future and that a negative result did not necessarily mean lack of protection against COVID-19.

### 2.5 Statistical analysis

Continuous variables like age were summarized with mean±SD, and categorical variables like gender were presented with count (%). Primary analyses, to evaluate diagnostic accuracy of the investigational kit, were reported with % for sensitivity and specificity. 95% confidence interval is specified for sensitivity and specificity. Positive predictive value specifies the proportions of true positive and negative predictive value species the proportions of true negative. In the vaccinated groups, true positives are the samples that tested positive using the study kit and the comparator kit and true negatives are the samples that tested negative using the study kit and the comparator kit. If the sample test positive with the gold standard but negative with the investigational kit, it was considered as false negative. If the sample test negative with the gold standard but positive with the investigational kit, it was considered as false positive. For pre-pandemic control samples, all the samples were considered as true negative as the samples were collected in 2018 before the emergence of SARS-CoV-2 and therefore antibodies against SARS-CoV-2 could not be present in these samples. So, in this group, any positives detected were considered as false positive and all negatives reported were considered true negatives. Sensitivity and specificity were calculated using the formulae:

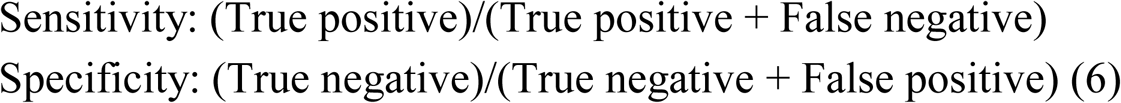

## 3 Results

Demographic data of the subjects in the vaccinated group (n=62) are presented in Table 1. The samples were collected at a median 62 (16-84) days after the second dose of vaccination in the cohort of the total vaccinated population. The mean age of this population was 36.5±11.58 years; of which 43.75% were male and 56.25% were female. Among this population, presence of co-morbidities such as hypertension, heart disease, and diabetes were 6.45%, 1.61%, and 4.84%, respectively. Demographic details and co-morbidities for the COVISHIELD- and Covaxin-vaccinated groups are presented separately in Table 1.

**Table 1:** Demographic details of the participants.

| **Characteristics** | **Combined vaccinated cohort** | **COVISHIELD -vaccinated group** | **Covaxin-vaccinated group** |
| --- | --- | --- | --- |
| **Age** | 36.65±11.58 | 36.56±12.20 | 36.73±11.09 |
| **Gender** |  |  |  |
| **Male** | 21(43.75) | 14 (43.75) | 7 (23.33) |
| **Female** | 41(56.25) | 18 (56.25) | 23 (76.67) |
| **Time span between 2nd dose of vaccine and blood collection (Days)** |  |  |  |
| **Average (Mean±SD)** | 58.45±19.61 | 62.33±21.62 | 55.46±17.63 |
| **Median (Min,Max)** | 62(16,84) | 70 (16, 84) | 58 (22, 83) |
| **Comorbidity, n(%)** |  |  |  |
| **Hypertension** | 4(6.45) | 2(6.25) | 2(6.67) |
| **heart disease** | 1(1.61) | 1(3.13) | 0 (0) |
| **Diabetes** | 3(4.84) | 1(3.13) | 2(6.67) |

For further clarity, we performed individual analysis of all four kits for both the vaccinated groups separately (Table 2). Analyses of Meril ABFind kit versus the other three kits as gold standard in the combined vaccinated cohort, COVISHIELD-vaccinated group and Covaxin-vaccinated group are shown in Table 3. In the combined vaccinated population, the Meril ABFind kit displayed 93.3% (CI, 89.83%-96.77%), 94.92% (CI, 91.88%-97.96%), and 90.3% (CI, 86.20%-94.4%) sensitivity in comparison to Abbott Quant, MERILISA, and GenScript cPass, respectively.

**Table 2:** Performance of different antibody detecting kits in COVISHIELD -vaccinated group and Covaxin-vaccinated group. (Abbott IgG II, Abbott SARS-CoV-2 IgG II Quant; GenScript, GenScript cPass SARS-CoV-2 neutralization antibody detection Kit; MERILISA, Covid Kawach MERILISA)

|  | COVISHIELD-vaccinated group |  |  |  | Covaxin-vaccinated group |  |  |  |
| --- | --- | --- | --- | --- | --- | --- | --- | --- |
| Results | Meril ABFind (n=32) | Abbott IgG II (n=32) | MERILIS A (n=32) | GenScript (n=32) | Meril ABFind (n=30) | Abbott IgG II (AU/mL ) (n=30) | MERILIS A (n=30) | GenScript (n=30) |
| Positive | 32 | 32 | 32 | 32 | 24 | 28 | 27 | 30 |
| Negative | 0 | 0 | 0 | 0 | 6 | 2 | 3 | 0 |

**Table 3:** Analysis of Meril ABFind kit versus the other three kits as gold standard in the combined vaccinated cohort, COVISHIELD-vaccinated group and Covaxin-vaccinated group. (Abbott IgG II, Abbott SARS-CoV-2 IgG II Quant; GenScript, GenScript cPass SARS-CoV-2 neutralization antibody detection Kit; MERILISA, Covid Kawach MERILISA)

|  | Combined vaccinated cohort |  |  | COVISHIELD-vaccinated group |  |  | Covaxin-vaccinated group |  |  |
| --- | --- | --- | --- | --- | --- | --- | --- | --- | --- |
| Parameters | Meril ABFind vs Abbott IgG II | Meril ABFind vs MERILISA | Meril ABFind vs GenScript | Meril ABFind vs Abbott IgG II | Meril ABFind vs MERILISA | Meril ABFind vs GenScript | Meril ABFind vs Abbott IgG II | Meril ABFind vs MERILISA | Meril ABFind vs GenScript |
| Sensitivity | 93.3% | 94.92% | 90.3% | 100% | 100% | 100% | 85.7% | 88.9% | 80% |
| Specificity | 100% | 100% | NA | NA | NA | NA | 100% | 100% | NA |
| Positive predictive value | 100% | 100% | 100% | 100% | 100% | 100% | 100% | 100% | NA |
| Negative predictive value | 33.3% | 50% | NA | NA | NA | NA | 100% | 100% | 100% |
| Confidence Interval for Sensitivity | 89.83% to 96.77% | 91.88% to 97.96% | 86.20% to 94.40% | NA | NA | NA | 71.71 to 99.71 | 76.33 to 100.00 | 64.00 to 96.00 |

In the COVISHIELD-vaccinated group, the results obtained with Meril ABFind kit were compared to the results obtained with other three kits considering them as “gold standard”. Samples from all the COVISHIELD recepients were positive by all four tests kits. The sensitivity of Meril ABFind kit in comparison to all the other three kits in this group was 100%. The positive predictive value calculated for Meril ABFind kit versus other three kits was also observed to be 100% In the Covaxin-vaccinated group, Meril ABFind kit displayed 85.7% (CI, 71.79%-99.71%), 88.9% (CI, 76.33%-100%), and 80% (CI, 64%-96%) sensitivity in comparison to Abbott Quant, MERILISA, and GenScript cPass, respectively. The specificity of the Meril ABFind kit in comparison to all the three kits was 100%.

The 40 pre-pandemic control samples tested with these four different kits showed different positive and negative results (Table 4). In the pre-pandemic controls, the Meril ABFind kit did not produce any false positive results. However, all the other three kits, Abbott Quant, MERILISA, and GenScript cPass did produce false positive results; 1, 4 and 5 false positives each, respectively. In this group, the specificity of Meril ABFind kit, Abbott Quant, MERILISA, and GenScript cPass were reported to be 100%, 97.5%, 90%, and 87.5% respectively.

**Table 4:**
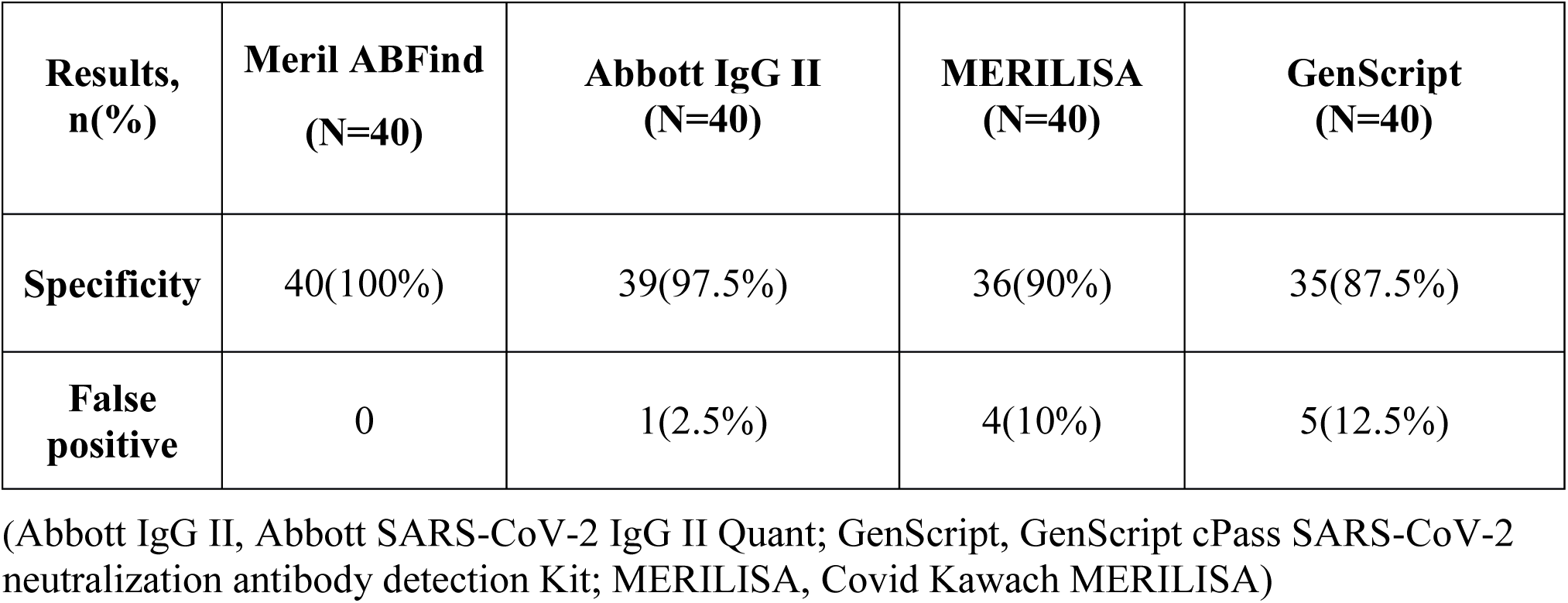
Performance of different antibody detecting kits in pre-pandemic control samples. (Abbott IgG II, Abbott SARS-CoV-2 IgG II Quant; GenScript, GenScript cPass SARS-CoV-2 neutralization antibody detection Kit; MERILISA, Covid Kawach MERILISA)

Quantitative antibody responses were significantly higher (p=0.01) after COVISHIELD vaccination (2889.36 ± 3572 AU/mL; CI 1651.72 to 4127) versus Covaxin vaccination (1054.13 ± 1360.01 AU/mL; CI 582.90 to 1525.35) detected by the Abbott Quant kit (Table 5).

**Table 5:**
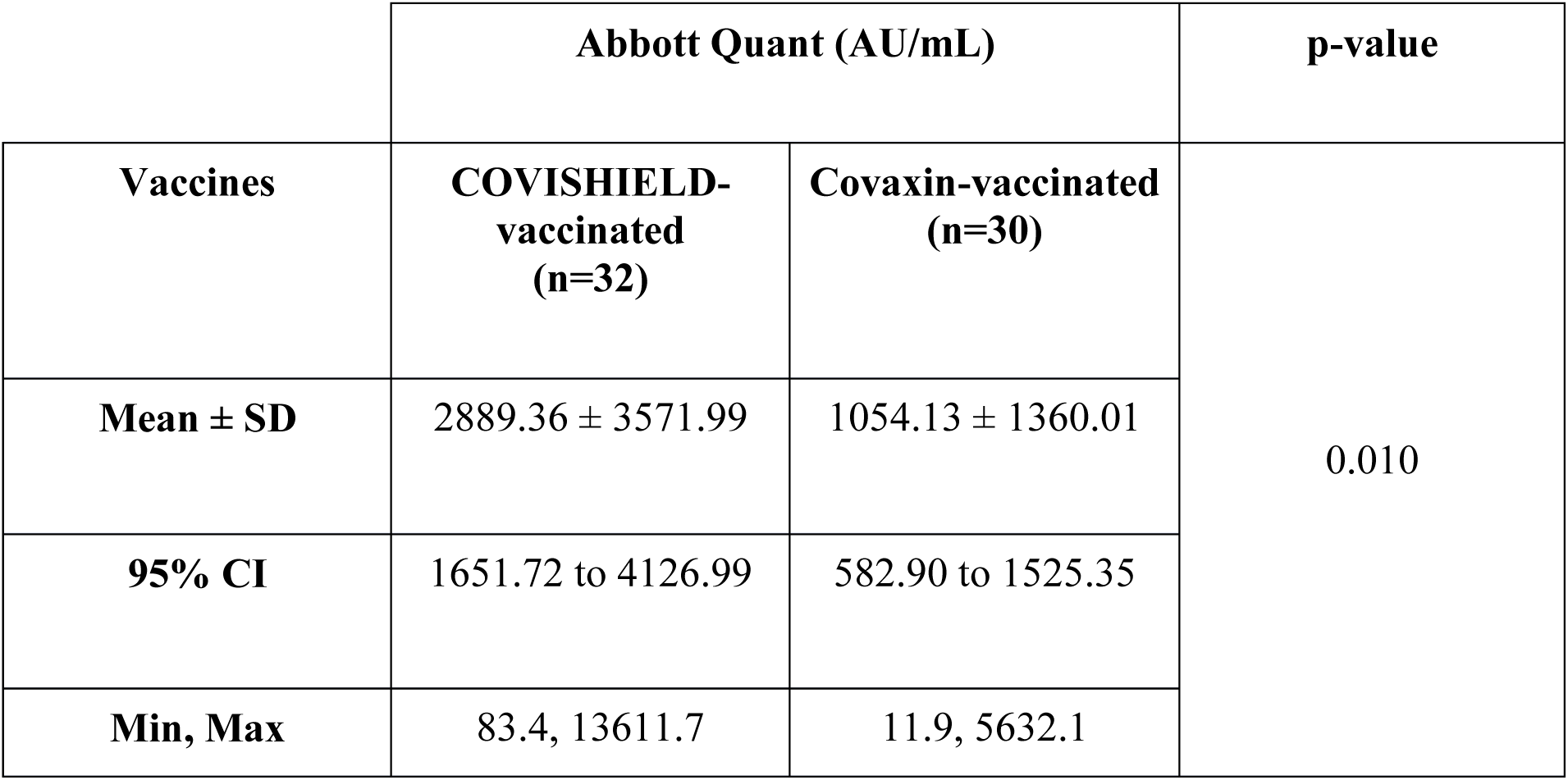
Quantitative determination of SARS-CoV-2 antibodies in COVISHIELD-vaccinated and Covaxin-vaccinated group by Abbott Quant.

## 4 Discussion

Previous studies have shown that neutralizing antibodies are targeted against spike protein, specifically the RBD and N-terminal domain of SARS-CoV-2 (8-10). Of monoclonal antibodies isolated from SARS-CoV-2 infected individuals, almost all those which potently neutralized SARS-CoV-2 *in vitro* were against RBD and a few against N-terminal domain (11).

Meril ABFind kit is based on detection of IgG antibodies against RBD in the specimens. This study evaluated the sensitivity and specificity of Meril ABFind kit in detecting COVID-19 antibodies in the samples collected from a diverse population. In this study, we had two groups of samples, one from COVID-19 vaccinated (either with COVISHIELD or Covaxin) group and the other from a pre-pandemic control group. The vaccinated group was further divided into a COVISHIELD-vaccinated group and a Covaxin-vaccinated group. The two vaccinated groups were comparable in terms of age and presence of pre-existing medical conditions (diabetes, hypertension, heart disease). The timing of sample collection from the second dose was also comparable in both the vaccinated groups.

The manufacturer claimed sensitivity and specificity of Abbott Quant were 99.35% and 99.6% (12), Covid Kawach MERILISA were approximately 93% and 100% (7) and GenScript cPass were 93.80% and 99.40%, respectively (13). Previous studies have also reported the sensitivity and specificity of the comparator kits. In one study, Abbott Quant kit demonstrated 96% sensitivity and 99.3% specificity in comparison to GenScript cPass (6). The Abbott Quant kit was also used in investigating the anti-spike IgG responses in health care workers following one or two doses of Pfizer-BioNTech or Oxford-Astrazenca vaccines (14). The GenScript cPass SARS-CoV-2 neutralization antibody detection kit is the first commercially available kit to detect neutralizing antibodies against SARS-CoV-2 RBD. One of the initial studies with this kit reported the sensitivity and specificity of this kit in the range of 77-100% and 95-100%, respectively, in comparison to plaque-reduction neutralization test and pseudo typed lentiviral neutralization assay (15). For our study, we have compared the performance of Meril ABFind kit with the other three commercially available kits and have not performed comparative analysis amongst the comparator kits. However, from the number of positive and negative cases detected, it can be inferred that the performance of all the kits used in this study were not identical.

The performance of the Meril ABFind kit was evaluated by comparing with the performance of the comparator kits viz. Abbott Quant, MERILISA, and GenScript cPass in both the groups. In the vaccinated population, the Meril ABFind kit displayed sensitivity of over 90% with all three comparators; 93.3% (CI, 89.83%-96.77%) with Abbott Quant, 94.92% (CI, 91.88%-97.96%) with MERILISA, and 90.3% (CI, 86.20%-94.4%) with GenScript cPass.

In the COVISHIELD-vaccinated group, in comparison to the other three kits the Meril ABFind kit displayed 100% sensitivity. As all the four kits have reported positive results and there were no negatives reported by any of them, calculating specificity of the kits was not valid for this group.

In the Covaxin-vaccinated group, the Meril ABFind kit had a sensitivity of 85.7% (CI, 71.79%-99.71%) with Abbott Quant, 88.9% (CI, 76.33%-100%) with MERILISA, and 80% (CI, 64%-96%) with GenScript cPass. The specificity was 100% against all three comparator tests.

A study in India comparing anti-Spike antibody responses following COVISHIELD and Covaxin found that seropositivity and geometric mean titers were higher following COVISHIELD vaccination compared to Covaxin vaccination (16). In our study, a significantly higher (p=0.01) level of anti-RBD IgG antibodies were detected in the COVISHIELD-vaccinated group versus the Covaxin-vaccinated group by the Abbott Quant kit. This may explain why Meril ABFind had a better sensitivity in the COVISHIELD group compared to the Covaxin group. Additionally, Meril ABFind does not detect antibodies against non-RBD epitopes. Covaxin is a whole virus inactivated vaccine and generates antibody responses against Spike and non-Spike antigens, while Meril ABFind only detects antibodies against the RBD. However, our study was not designed to compare the seropositivity between COVISHIELD- and Covaxin-vaccinated individuals. Based on our study, we cannot comment on the immunogenicity or efficacy of COVISHIELD compared with Covaxin.

The control samples were collected before the pandemic, and in this group the results of all four kits differed. It was deemed impossible for these samples to contain antibodies against specific SARS-CoV-2 antigens as these samples were collected in 2018. Any positives in these samples would necessarily be false positives due to cross reactivity with antibodies against other endemic coronaviruses or other non-coronavirus pathogens. These samples were collected from patients with an acute febrile illness which included dengue and leptospirosis but may have also included other infectious diseases which may have resulted in cross reactivity and false positives.

In the pre-pandemic control samples, all positive results by Abbott Quant, MERILISA, and GenScript cPass were classified as false positive results as it was not possible for these samples to contain antibodies against SARS-CoV-2. The Meril ABFind kit did not produce a single false positive in the pre-pandemic controls, and therefore in this cohort, Meril ABFind kit was 100% specific, which was better than Abbott Quant IgG II, MERILISA, and GenScript cPass (Figure 1).

**Figure 1:**
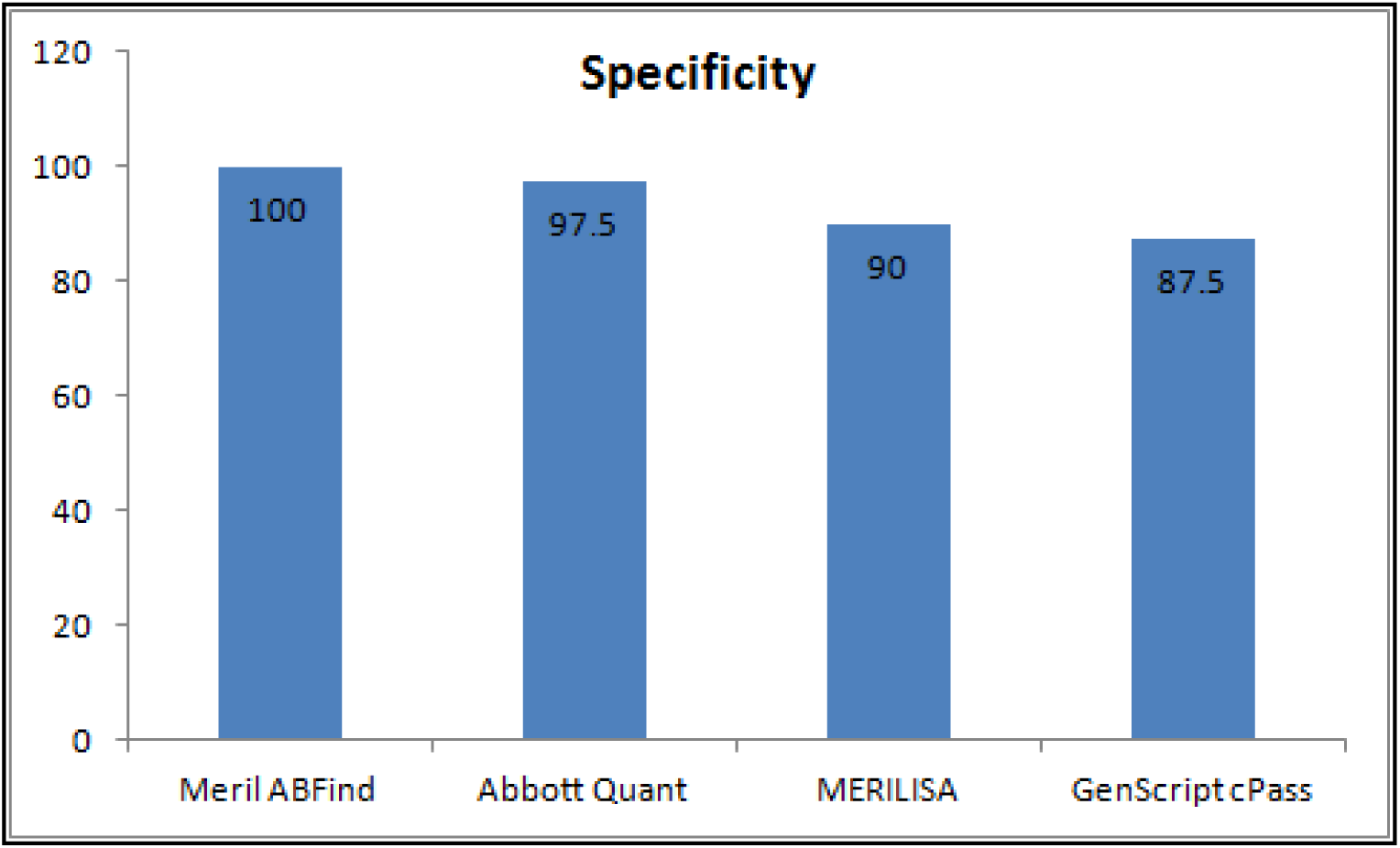
Specificity of the four kits in the pre-pandemic control group.

Interestingly, Abbott Quant, MERILISA, and GenScript cPass detected 2.5%, 10%, and 12.5% false positives, respectively. As all three of these kits (Abbott Quant, MERILISA, and GenScript cPass kits) produced false-positive results in the pre-pandemic samples, it is possible that they produced some false positives in the samples from the Covaxin-vaccinated subjects as well. It is possible that Meril ABFind displayed a higher sensitivity in the Covaxin group while the comparator kits (Abbott Quant, MERILISA, and GenScript cPass) displayed specificity lower than reported in literature. It would be prudent to assess these assays in a large cohort of pre-pandemic samples from India to ensure absence of cross-reactivity against non-SARS-CoV-2 coronaviruses or other pathogens.

Currently, many diagnostic kits which were developed to detect antibodies against SARS-CoV-2 in specimens collected from infected and vaccinated individuals against COVID-19 are commercially available. The rapid lateral flow serological assays were the first commercially available serological assays, but their poor sensitivity and specificity made them unsuitable for use (11). Since then numerous laboratory-based assays, for example CLIA & ELISA have been introduced in India with reported high sensitivity and specificity. However, there is a lack of laboratory infrastructure, technical expertise in rural India for such assays. Serological assays have demonstrated great value in serological studies to understand the percentage of the population that has COVID-19 antibodies. Such studies will continue to be relevant in the future. Serology has also been used to guide clinical decision making. In the UK, NHS guidelines recommend offering a combination of casirivimab and imdevimab to people aged 12 and over in hospital with COVID-19 who have no detectable SARS-CoV-2 antibodies (seronegative) (3). This treatment works best when administered early in disease course and accurate point of care serological tests may offer value in this setting. While serology is not currently used to guide decisions on vaccination, it is possible that in the future decisions on the need for additional doses of COVID-19 vaccines or boosters will be based on serology. Therefore, an accurate point of care serological test may offer value in future serological studies, clinical decision making and vaccination programs. A rapid and accurate serological assay that can detect post vaccination COVID-19 neutralizing antibodies would be especially useful in areas where laboratory facilities are not available.

In this study, the Meril ABFind kit showed excellent sensitivity and specificity in the COVISHIELD- and Covaxin-vaccinated groups. Notably, Meril ABFind kit was the only assay in this study that had a zero false positivity rate and 100% specificity in the pre-pandemic samples, while all three comparator kits had some false positives. This suggests the conventional thought process that rapid point of care tests are necessarily less accurate or specific than laboratory based assays may not always hold true, at least as concerns SARS-CoV-2 serological tests.

Our study also highlights the need to investigate laboratory based assays for cross reactivity in the Indian population. If we had not used pre-pandemic samples as a control, we would have missed the possibility that Abbott Quant, MERILISA, and GenScript cPass had a false positivity rate as high as 2.5%, 10% and 12.5%. While it is important to have sensitive assays that can correctly detect all individuals who have an adaptive humoral response to SARS-CoV-2, it is equally important to ensure that an assay is specific and doesn’t incorrectly label those who do not have antibodies as being sero-positive, particularly for neutralizing antibodies against SARS-COV-2. False positive results in serological studies may result in over-estimating the population that has antibodies from infection and vaccination, and this may have important implications on policy decisions. A false positive result may disqualify an individual from a therapeutic like a monoclonal antibody cocktail when it is dependant on serological status.

Our study shows that point of care serological tests can be a suitable alternative to laboratory based assays. Meril ABFind kit has an excellent sensitivity and specificity in comparison to commercially available laboratory assays and holds promise in accurate identification of SARS-CoV-2 anti-RBD neutralizing antibodies. In our study, Meril ABFind kit was the only assay that didn’t produce false positives. The results of the control pre-pandemic samples show that the Meril ABFind kit had the highest specificity in pre-pandemic control samples. However, larger studies with larger sample size are warranted to provide conclusive evidence of comparative performance of all the kits used in this study.

## Data Availability

All data produced in the present work are contained in the manuscript

## 5 Conflict of Interest

The authors declare that the research was conducted in the absence of any commercial or financial relationships that could be construed as a potential conflict of interest.

## 6 Author Contributions

JS conceptualized the study. SA, NC and HG collected and compiled data. JS, SA, NC and HG drafted the manuscript, prepared tables and figures. All authors contributed to data interpretation, writing, critically reviewing, revising, and approving the final manuscript for submission.

## 7 Funding

This work was supported by the Municipal Corporation of Greater Mumbai’s funding for Kasturba Hospital’s COVID-19 diagnostic laboratory. GenScript cPass SARS-CoV-2 neutralization antibody detection kits, COVID Kawach MERILISA kits and Meril ABFind kits were provided by Meril Diagnostic Pvt Limited.

## 8 Acknowledgments

The authors acknowledge and thank the Municipal Corporation of Greater Mumbai for their support through the study. We thank Meril Diagnostic Pvt. Limited for providing test kits to conduct the study. We thank the health workers who participated in this study.

## 9 Data Availability Statement

The original contributions presented in the study are included in the article. Further inquiries can be directed to the corresponding author.

## Notes

### Competing Interest Statement

The authors have declared no competing interest.

### Funding Statement

This work was supported by the Municipal Corporation of Greater Mumbais funding for Kasturba Hospital's COVID-19 diagnostic laboratory. Test kits were provided by Meril Diagnostic Pvt Limited.

### Author Declarations

The studies involving human participants were reviewed and approved by the Institutional Review Board of Kasturba Hospital of Infectious Diseases; IRB number 05/2021. The patients/participants provided their written informed consent to participate in this study. All collection, processing of samples and archiving of results were performed under approval from Institutional Review Board.

